# The Psychological Footprint of Unruptured Intracranial Aneurysm Discovery

**DOI:** 10.64898/2026.08.25.26361377

**Authors:** Daniela B. Renedo, Huanwen “Alvin” Chen, Kevin N. Sheth, Dheeraj Gandhi, Ajay Malhotra, Charles C. Matouk

## Abstract

**Background:** Unruptured intracranial aneurysms (UIAs) are increasingly identified incidentally, and management balances rupture risk against treatment risk. UIA diagnosis has been linked to psychological distress, but psychotropic medication initiation after UIA discovery has not been compared across the full UIA management spectrum.

**Methods:** We conducted a retrospective cohort study using IBM MarketScan claims (CCAE, MDCD, and MDCR; 2009–2023) among adults with a UIA diagnosis, continuous enrollment for 365 days before and after the index date, and no SAH/rupture on or before the index date. We compared the prevalence of 6 mental-health diagnoses before versus after UIA discovery and used adjusted logistic regression to examine psychotropic medication initiation within 365 days by management strategy (untreated observation as the reference).

**Results:** Among 54,945 patients (untreated, 78.5%; endovascular, 11.3%; clipping, 3.0%; other/uncertain, 7.2%), prevalence of every mental-health diagnosis was higher after UIA discovery, most for depression (+4.6 percentage points) and anxiety (+4.5 points). Medication initiation was most common for benzodiazepines (8.7%). Endovascular treatment was associated with higher adjusted odds of benzodiazepine (aOR, 1.21), SSRI (aOR, 1.20), and sedative-hypnotic (aOR, 1.25) initiation.Surgical clipping demonstrated the broadest association, with higher odds across 5 of 6 classes, including benzodiazepines (aOR, 1.71) and sedative-hypnotics (aOR, 1.86). Benzodiazepines had the lowest 1-year persistence (10.5%) despite being the most commonly initiated class. Findings were consistent across sensitivity analyses, with the exception of the increase in panic disorder, which was no longer observed after applying a 30-day post-index lag.

**Conclusions:** Mental-health diagnoses and psychotropic medication initiation increased after UIA discovery, and medication initiation was most pronounced among patients treated with surgical clipping. These findings support psychological assessment as part of aneurysm management regardless of strategy.

## Introduction

Unruptured intracranial aneurysms (UIAs) affect approximately 1%–2% of adults and are increasingly identified as incidental findings on cross-sectional neuroimaging obtained for unrelated indications.^1,2^ Management requires weighing a generally low annual rupture risk against the risks of preventive treatment, and current guidelines support either long-term observation or repair by endovascular treatment or surgical clipping depending on aneurysm and patient characteristics.^3,4^

Although rupture risk and procedural safety have a central role in the decision-making process, a diagnosis of UIA also carries a psychological burden independent of whether the aneurysm is treated. Patients often describe living with an aneurysm as carrying a “ticking time bomb,” and case-control and cross-sectional studies have linked UIA diagnosis with elevated anxiety and reduced quality of life even without rupture.^1,2,5^ Two recent large registry studies reinforce this burden: one found a higher long-term incidence of mental-illness diagnoses among untreated UIA patients compared with matched non-UIA controls, and another linked post-diagnostic anxiety and depression to a reduced likelihood of preventive treatment and worse rupture and mortality outcomes.^5,6^

This literature has largely compared patients with UIAs with external controls or examined mental health as a predictor of subsequent treatment choice, typically within a single-country UIA population (untreated or general). Little is known about the pharmacologic burden of UIA discovery—specifically, new psychotropic medication initiation and persistence, claims-based measures of treatment-seeking that extend beyond diagnosis codes alone—or whether this burden differs across the full management spectrum of observation, endovascular treatment, and surgical clipping within the same cohort. To our knowledge, this question has not been examined in a large US-insured population.

We used a multistate US administrative claims database to examine (1) whether the prevalence of 6 common mental-health diagnoses changed from the year before to the year after UIA discovery and (2) whether initiation and persistence of psychotropic medications during the year after discovery differed by management strategy (observation alone, endovascular treatment, or surgical clipping).

## Methods

### Study Design and Data Source

We conducted a retrospective cohort study using deidentified claims from the IBM MarketScan Commercial Claims and Encounters (CCAE), Multi-State Medicaid (MDCD), and Medicare Supplemental and Coordination of Benefits (MDCR) databases, which include enrollment, inpatient, outpatient, and outpatient pharmacy claims. Index dates ranged from 2009 through 2023.^7,8^

### Study Population

Patients with an unruptured intracranial aneurysm (UIA) were identified from a previously constructed MarketScan cohort using ICD-9-CM code 437.3 and ICD-10-CM code I67.1 and were restricted to those aged 18 years or older with a valid enrollee identifier and index date. The index date was defined as the date of the first observed claim carrying a UIA diagnosis code in the source cohort; because claims cannot establish the first-ever clinical diagnosis, “discovery” refers operationally to this first observed claim. Eligible patients had continuous enrollment for 365 days before and after the index date and no diagnosis of subarachnoid hemorrhage (SAH) or aneurysm rupture on or before the index date. These criteria yielded a final analytic cohort of 54,945 patients (Figure 1). Diagnosis and procedure code definitions for cohort identification, the SAH/rupture exclusion, and all covariates are provided in **Supplementary Table S1**.

**Figure 1.**
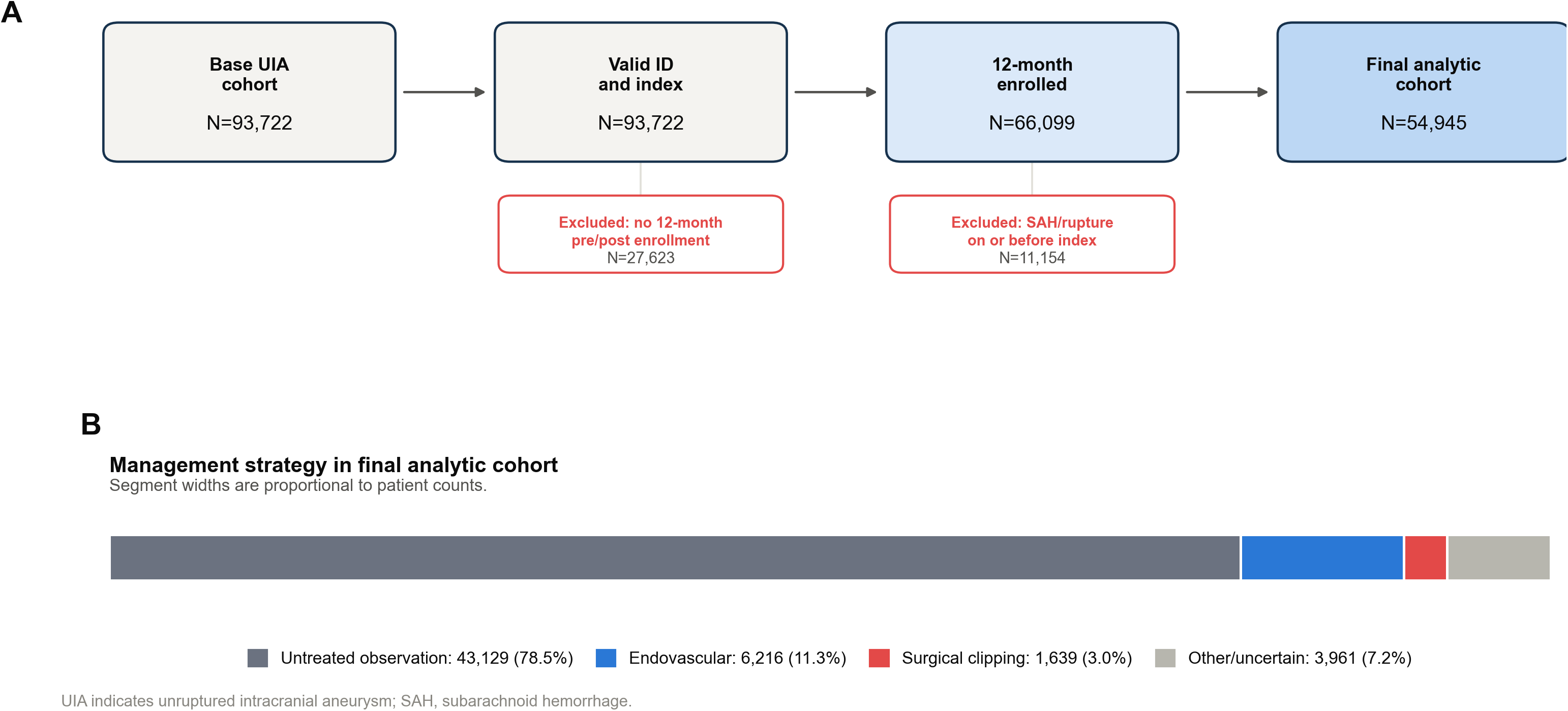
Study cohort selection and management classification.

### Management Strategy

Patients were classified from procedure and diagnosis claims into 4 mutually exclusive groups: surgical clipping, endovascular treatment, other/uncertain, or untreated observation (no repair code identified). Code definitions are provided in **Supplementary Table S1**. The other/uncertain group was included in descriptive analyses only and excluded from adjusted models.

### Mental-Health Outcomes

We assessed 6 mental-health diagnoses: depression, anxiety, insomnia/sleep disorder, adjustment disorder, posttraumatic stress disorder, and panic disorder, identified from inpatient, outpatient, and facility claims (code definitions, Supplementary Table S1). These diagnoses were selected to capture the clinical components of anxiety- and stress-related morbidity most consistently reported in prior UIA cohorts.^9^ For each diagnosis, we compared the proportion of patients with at least 1 qualifying claim during the 365-day baseline period with the proportion during the 365 days after the index date.

### Psychotropic Medication Initiation and Persistence

Outpatient pharmacy claims were used to identify fills of 6 psychotropic medication classes: benzodiazepines, selective serotonin reuptake inhibitors (SSRIs), serotonin-norepinephrine reuptake inhibitors (SNRIs), other antidepressants, non-benzodiazepine anxiolytics, and sedative-hypnotics, mapped by generic name and National Drug Code (Supplementary Table S1). This 6-class structure follows new-user designs used in prior claims-based studies of psychotropic initiation after a health-related or life stressor,^8,9^ with medication classes subdivided by pharmacologic mechanism and clinical use given their distinct safety profiles, particularly in older adults.^10,11^ For each class, eligible new users were patients with no fill during the 365-day baseline period. Initiation was defined as the first qualifying fill within 365 days after the index date. Among new initiators, persistence was defined using cumulative days supplied: at least 60 days by day 90, at least 90 days by day 180, and at least 180 days by day 365. We also summarized total days supplied and, for benzodiazepines, long-term exposure (at least 120 days supplied within 365 days).

### Covariates

Adjusted models included age, sex, MarketScan database, Charlson comorbidity score (derived from 17 baseline diagnostic categories), baseline migraine/headache diagnosis, and log-transformed baseline medical claim count. Diagnosis codes used to define covariates are provided in **Supplementary Table S1**.

### Statistical Analysis

Categorical variables are reported as counts and percentages, and continuous variables as means with SD. For each medication class, we fit multivariable logistic regression models among class-specific eligible new users, with initiation within 365 days as the outcome and management strategy (endovascular treatment or surgical clipping vs untreated observation as reference) as the exposure, adjusted for the covariates above. Cell counts <11 were suppressed per the data use agreement. Pre-versus post-index mental-health diagnosis prevalence was compared within patients using the exact McNemar test on discordant pairs; we report the paired risk difference with 95% CI and the within-person odds ratio. Adjusted management-strategy models were also re-fit with index year included as a covariate. All analyses were performed with R version 4.5.2.

### Sensitivity Analyses

We performed 2 prespecified sensitivity analyses: (1) a 30-day lag analysis excluding the first 30 days after the index date to reduce detection bias, and (2) a redefinition of the SAH/rupture exclusion to claims occurring during the 365-day baseline period only, rather than at any time before the index date, with all primary analyses repeated in the resulting cohort.

### Ethics and Data Availability

The data supporting the findings of this study were obtained under license from the Merative™ MarketScan® Research Databases. Due to licensing agreements and third-party commercial restrictions, these data are not publicly available and cannot be redistributed by the investigators. Researchers interested in accessing the raw dataset must apply directly for a license through Merative (). Under institutional policy, analyses of these deidentified data did not constitute human-subjects research; therefore, institutional review board approval and informed consent were not required. DR had full access to all data used in the study and takes responsibility for the integrity of the data and the accuracy of the data analysis.

This study was reported in accordance with the STROBE statement and the RECORD extension for routinely collected health data.

## Results

### Study Cohort and Baseline Characteristics

Among 93,722 patients with a UIA diagnosis across CCAE, MDCD, and MDCR, 66,099 (70.5%) had continuous enrollment for 365 days before and after the index date. After excluding 11,154 patients with SAH or aneurysm rupture on or before the index date, 54,945 patients composed the final analytic cohort **(Figure 1)**. Management strategy was observation alone in 43,129 (78.5%), endovascular treatment in 6,216 (11.3%), surgical clipping in 1,639 (3.0%), and other/uncertain in 3,961 (7.2%) **(Table 1)**. Mean age was 59.3 years and 71.8% of patients were women; patients who underwent clipping or endovascular treatment were younger, on average, and had a higher prevalence of baseline migraine/headache than patients managed with observation alone **(Table 1)**.

**Table 1.**
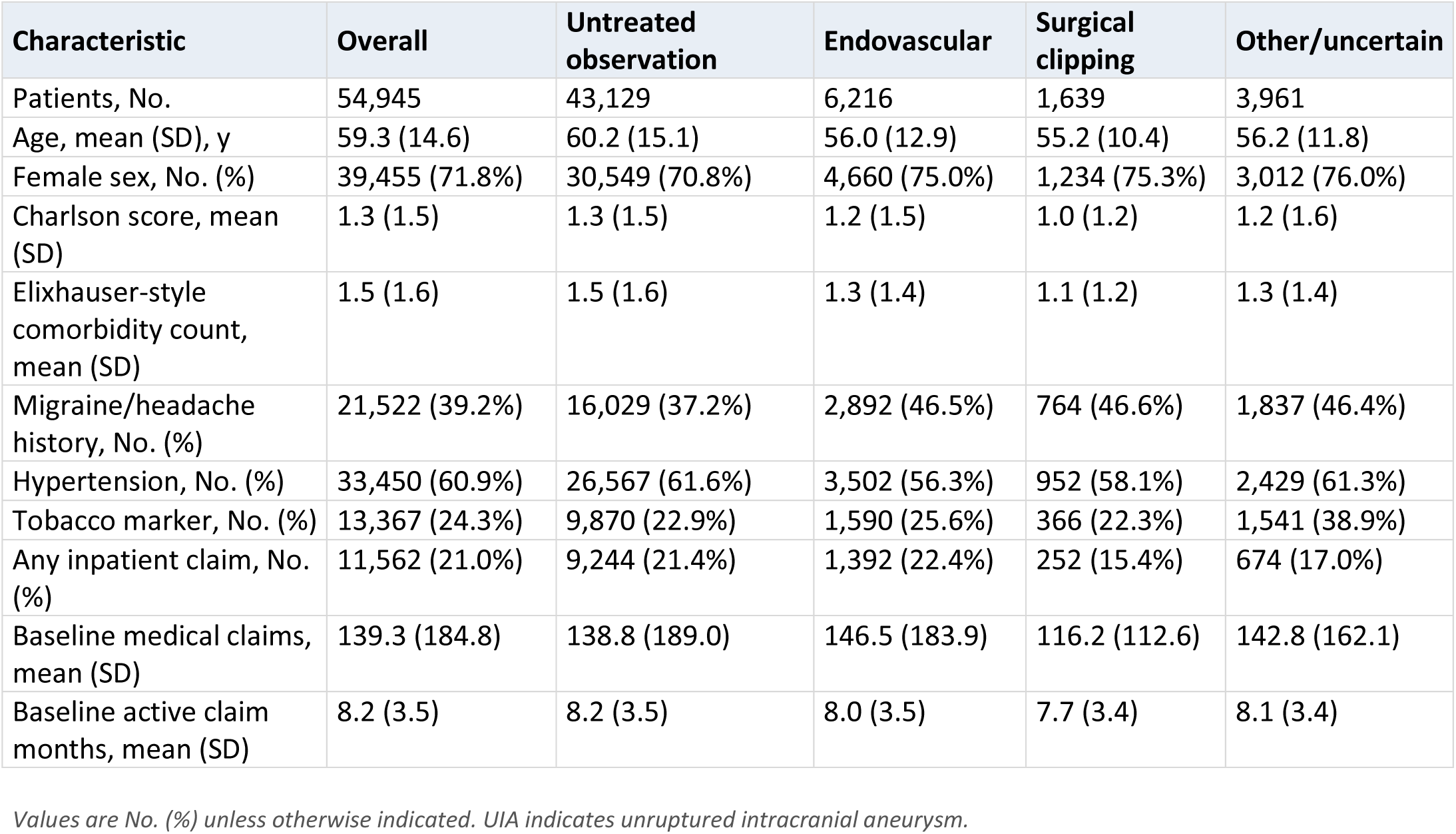
Baseline characteristics according to UIA management strategy.

| Characteristic | Overall | Untreated observation | Endovascular | Surgical clipping | Other/uncertain |
| --- | --- | --- | --- | --- | --- |
| Patients, No. | 54,945 | 43,129 | 6,216 | 1,639 | 3,961 |
| Age, mean (SD), y | 59.3 (14.6) | 60.2 (15.1) | 56.0 (12.9) | 55.2 (10.4) | 56.2 (11.8) |
| Female sex, No. (%) | 39,455 (71.8%) | 30,549 (70.8%) | 4,660 (75.0%) | 1,234 (75.3%) | 3,012 (76.0%) |
| Charlson score, mean (SD) | 1.3 (1.5) | 1.3 (1.5) | 1.2 (1.5) | 1.0 (1.2) | 1.2 (1.6) |
| Elixhauser-style comorbidity count, mean (SD) | 1.5 (1.6) | 1.5 (1.6) | 1.3 (1.4) | 1.1 (1.2) | 1.3 (1.4) |
| Migraine/headache history, No. (%) | 21,522 (39.2%) | 16,029 (37.2%) | 2,892 (46.5%) | 764 (46.6%) | 1,837 (46.4%) |
| Hypertension, No. (%) | 33,450 (60.9%) | 26,567 (61.6%) | 3,502 (56.3%) | 952 (58.1%) | 2,429 (61.3%) |
| Tobacco marker, No. (%) | 13,367 (24.3%) | 9,870 (22.9%) | 1,590 (25.6%) | 366 (22.3%) | 1,541 (38.9%) |
| Any inpatient claim, No. (%) | 11,562 (21.0%) | 9,244 (21.4%) | 1,392 (22.4%) | 252 (15.4%) | 674 (17.0%) |
| Baseline medical claims, mean (SD) | 139.3 (184.8) | 138.8 (189.0) | 146.5 (183.9) | 116.2 (112.6) | 142.8 (162.1) |
| Baseline active claim months, mean (SD) | 8.2 (3.5) | 8.2 (3.5) | 8.0 (3.5) | 7.7 (3.4) | 8.1 (3.4) |

Values are No. (%) unless otherwise indicated. UIA indicates unruptured intracranial aneurysm.

### Mental-Health Diagnoses Following UIA Discovery

The prevalence of every assessed mental-health diagnosis was higher during the 365 days after UIA discovery than during the 365 days before the index date (Figure 2): depression (+4.6 percentage points), anxiety (+4.5), insomnia/sleep disorder (+3.8), adjustment disorder (+1.5), PTSD (+0.5), and panic disorder (+0.3). Among patients with a qualifying post-index diagnosis, median time to first diagnosis ranged from 53 days (depression and anxiety) to 79 days (panic disorder). Paired within-person comparisons confirmed these increases: depression, paired risk difference +4.6 percentage points (95% CI, 4.3–5.0), within-person OR 1.82 (95% CI, 1.74–1.90); anxiety, +4.5 points (95% CI, 4.2–4.8), OR 1.69 (95% CI, 1.62–1.76); insomnia/sleep disorder, +3.8 points (95% CI, 3.4–4.1), OR 1.59 (95% CI, 1.52–1.66); adjustment disorder, +1.5 points (95% CI, 1.3–1.7), OR 1.49 (95% CI, 1.40–1.59); PTSD, +0.5 points (95% CI, 0.3–0.6), OR 1.59 (95% CI, 1.41–1.80); and panic disorder, +0.3 points (95% CI, 0.1–0.4), OR 1.27 (95% CI, 1.13–1.42) **(Supplementary Table S2)**.

**Figure 2.**
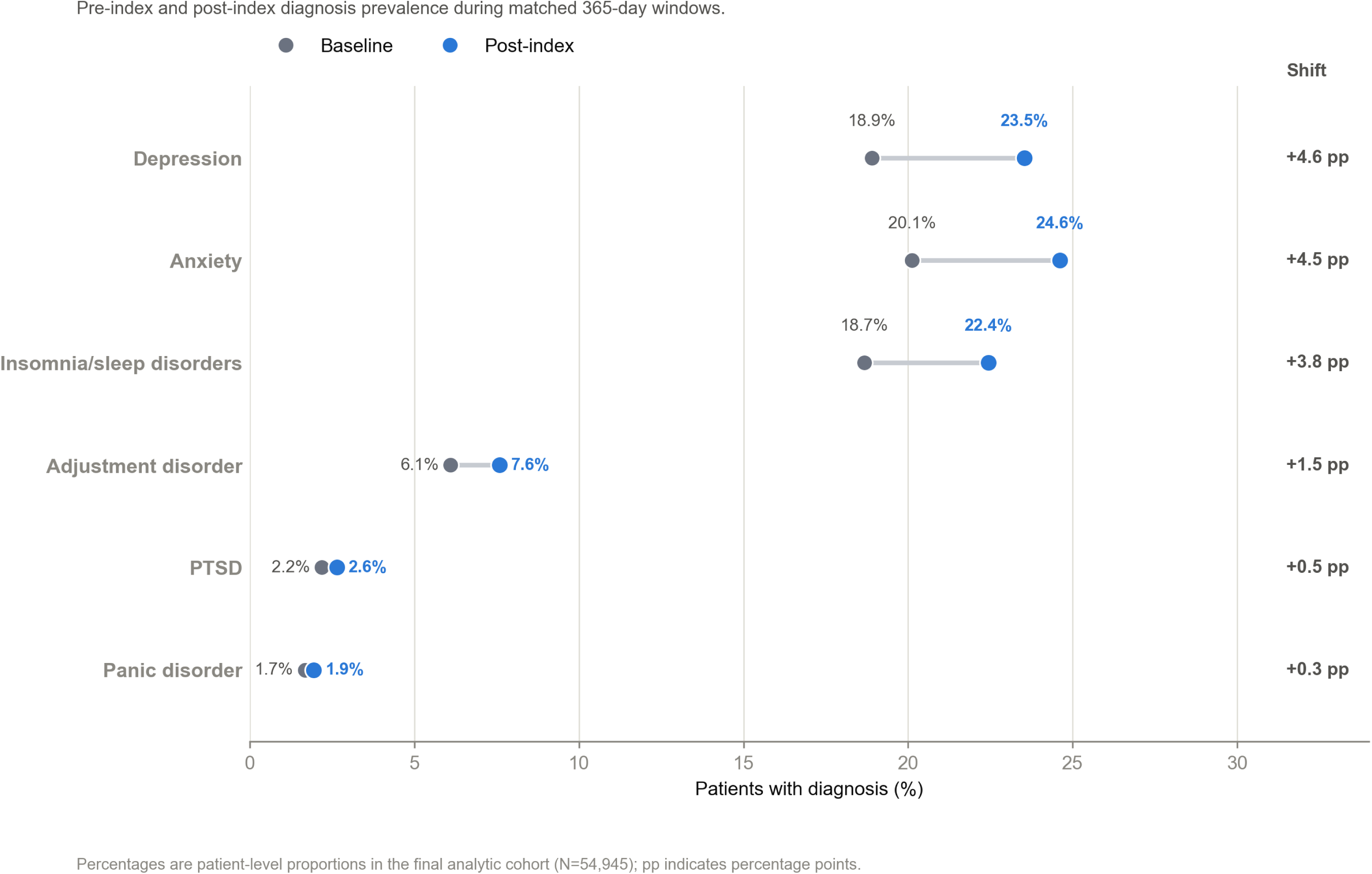
Mental-health diagnoses before and after UIA discovery.

Among patients without a baseline diagnosis, 1-year cumulative incidence was 12.7% (95% CI, 12.4–13.0) for depression, 13.8% (95% CI, 13.5–14.2) for anxiety, and 12.5% (95% CI, 12.2–12.8) for insomnia/sleep disorder.

### New Psychotropic Medication Initiation

Among class-specific eligible new users, initiation within 365 days was most common for benzodiazepines (3,738/42,839, 8.7%), other antidepressants (3,582/46,115, 7.8%), and SSRIs (3,443/44,783, 7.7%), and less common for non-benzodiazepine anxiolytics (3.7%), sedative-hypnotics (3.2%), and SNRIs (2.9%) **(Table 2)**. Median time to initiation ranged from 105 days (benzodiazepines) to 156 days (SNRIs).

**Table 2.** New psychotropic medication initiation during the year following UIA discovery.

| Medication class | Eligible new-user population, No. | Initiated within 365 d, No. | Initiation, % (95% CI) | Initiated by 30 d, No. | Initiated by 90 d, No. | Initiated by 180 d, No. | Median days to initiation |
| --- | --- | --- | --- | --- | --- | --- | --- |
| Benzodiazepines | 42,839 | 3,738 | 8.73 (8.46–9.00) | 946 | 1,730 | 2,485 | 105 |
| SSRIs | 44,783 | 3,443 | 7.69 (7.45–7.94) | 651 | 1,450 | 2,244 | 119 |
| SNRIs | 50,936 | 1,465 | 2.88 (2.73–3.02) | 182 | 465 | 814 | 156 |
| Other antidepressants | 46,115 | 3,582 | 7.77 (7.53–8.02) | 730 | 1,513 | 2,344 | 120 |
| Non-benzodiazepine anxiolytics | 51,170 | 1,911 | 3.73 (3.57–3.90) | 348 | 741 | 1,185 | 129 |
| Sedative-hypnotics | 50,348 | 1,608 | 3.19 (3.04–3.35) | 307 | 648 | 1,035 | 121 |

### Association Between Management Strategy and Medication Initiation

In unadjusted comparisons, initiation of all 6 medication classes was more common in the treated or uncertain group than among patients managed with observation alone, with unadjusted odds ratios ranging from 1.10 (95% CI, 0.99–1.23) for non-benzodiazepine anxiolytics to 1.41 (95% CI, 1.30–1.52) for benzodiazepines **(Supplementary Table S3)**.

In adjusted models, endovascular treatment (vs untreated observation) was associated with higher odds of benzodiazepine (aOR, 1.21; 95% CI, 1.09–1.34), SSRI (aOR, 1.20; 95% CI, 1.08–1.34), and sedative-hypnotic initiation (aOR, 1.25; 95% CI, 1.08–1.45) and with lower odds of non-benzodiazepine anxiolytic initiation (aOR, 0.85; 95% CI,0.73– 0.99). The latter association was attenuated in the SAH-window sensitivity analysis (P=.09). Surgical clipping was associated with higher odds of initiation across 5 of 6 medication classes: benzodiazepines (aOR, 1.71; 95% CI, 1.44–2.02), SSRIs (aOR, 1.57; 95% CI, 1.31–1.88), other antidepressants (aOR, 1.39; 95% CI, 1.16–1.67), sedative-hypnotics (aOR, 1.86; 95% CI, 1.46–2.36), and SNRIs (aOR, 1.35; 95% CI, 1.03–1.76) (Figure 3; Supplementary Table S4). Clipping was not associated with non-benzodiazepine anxiolytic initiation (aOR, 1.01; 95% CI, 0.77–1.34). After adjustment for index year, all 5 clipping associations remained significant. The benzodiazepine and SSRI associations with endovascular treatment were essentially unchanged, but the endovascular/sedative-hypnotic association was no longer significant (aOR, 1.12; 95% CI, 0.96–1.31) **(Supplementary Table S5)**.

**Figure 3.**
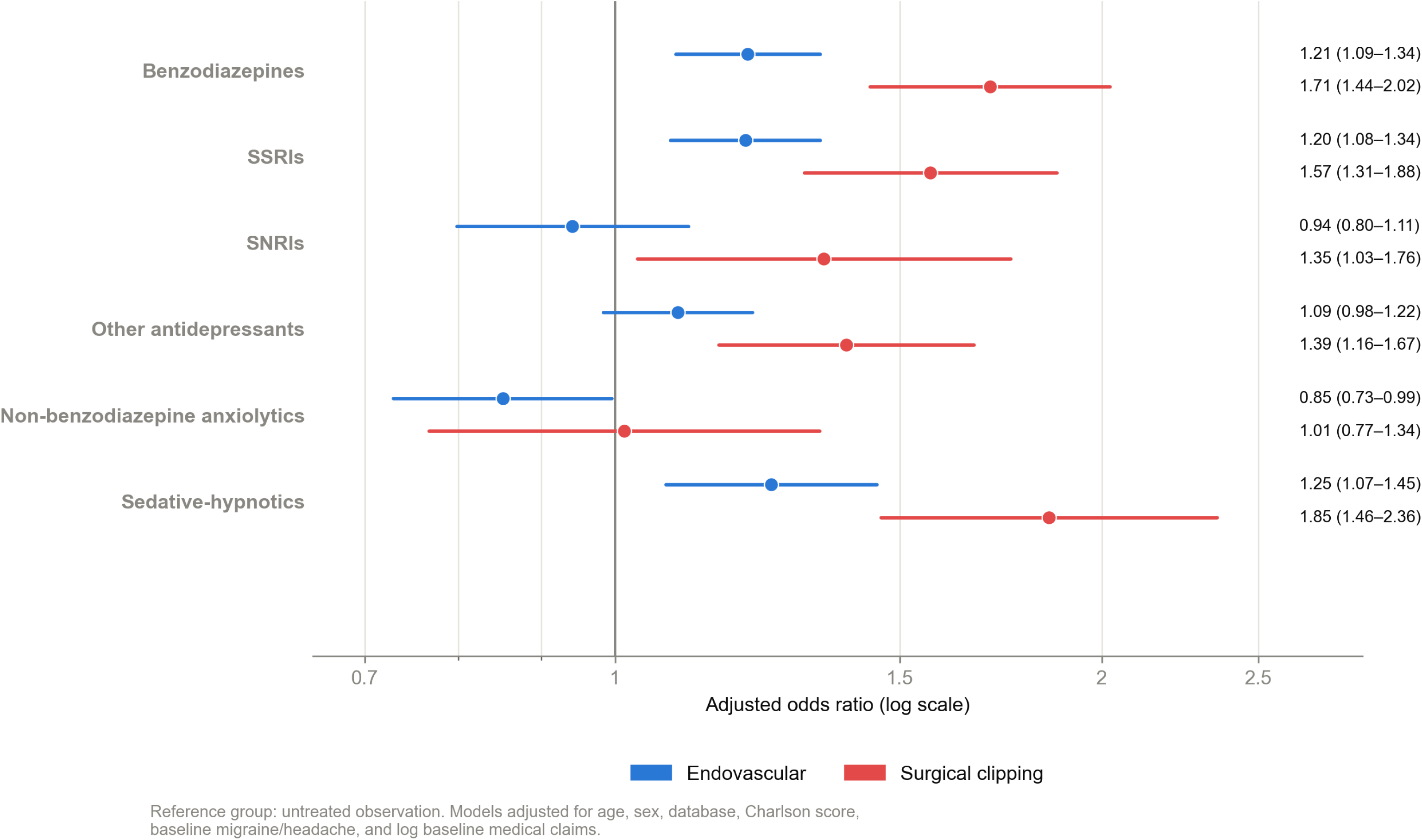
Adjusted association between management strategy and psychotropic medication initiation.

### Persistence of Newly Initiated Medications

At 365 days, persistence among new initiators was highest for SSRIs (37.3%; 95% CI, 35.7–38.9), SNRIs (31.3%; 95% CI, 28.9–33.7), and other antidepressants (27.6%; 95% CI, 26.2–29.1) and lowest for sedative-hypnotics (17.1%; 95% CI, 15.3–19.0), non-benzodiazepine anxiolytics (16.0%; 95% CI, 14.4–17.7), and benzodiazepines (10.5%; 95% CI, 9.6–11.5) (Table 3). Median total days supplied was 100 days (IQR, 30–240) for SSRI initiators versus 24 days (IQR, 5–60) for benzodiazepine initiators, consistent with predominantly short-term or situational benzodiazepine use. Among 3,738 benzodiazepine initiators, 590 (15.8%) met a long-term exposure threshold of at least 120 days supplied within 365 days.

**Table 3.** Persistence of newly initiated psychotropic medications.

| Medication class | New initiators, No. | Persistent at 90 d, No. (%) | Persistent at 180 d, No. (%) | Persistent at 365 d, % (95% CI) | Median total days supplied | Long-term benzodiazepine exposure, 120 d supplied/365 d |
| --- | --- | --- | --- | --- | --- | --- |
| Benzodiazepines | 3,738 | 281 (7.5%) | 392 (10.5%) | 10.52 (9.58–11.54) | 24 (5–60) | 590 (15.8%) |
| SSRIs | 3,443 | 819 (23.8%) | 1,188 (34.5%) | 37.27 (35.67–38.90) | 100 (30–240) | NA |
| SNRIs | 1,465 | 237 (16.2%) | 385 (26.3%) | 31.26 (28.94–33.68) | 90 (30–186) | NA |
| Other antidepressants | 3,582 | 705 (19.7%) | 991 (27.7%) | 27.61 (26.17–29.09) | 90 (30–180) | NA |
| Non-benzodiazepine anxiolytics | 1,911 | 229 (12.0%) | 307 (16.1%) | 15.96 (14.39–17.67) | 30 (15–100) | NA |
| Sedative-hypnotics | 1,608 | 232 (14.4%) | 294 (18.3%) | 17.10 (15.34–19.02) | 60 (30–120) | NA |

*Persistence thresholds: 90 d, at least 60 days supplied by day 90; 180 d, at least 90 days supplied by day 180; 365 d, at least 180 days supplied by day 365.*

### Sensitivity Analyses

Findings were consistent across both sensitivity analyses. In the 30-day lag analysis, post-index increases in depression, anxiety, and insomnia/sleep disorder were attenuated but remained present (Supplementary Table S6). The increase in panic disorder was no longer observed after the 30-day lag (paired risk difference, −0.02 percentage points; 95% CI, −0.1 to +0.1; P=.82), consistent with panic symptoms arising during the diagnostic evaluation that identified the aneurysm rather than after its discovery. Redefining the SAH/rupture exclusion window to the 365-day baseline period added 1,109 patients to the analytic cohort (54,945 to 56,054). The largest resulting change in a mental-health prevalence shift was 0.08 percentage points, and the largest change in a medication-initiation rate was 0.07 percentage points. No adjusted odds-ratio direction changed. The only significance-threshold change was the non-benzodiazepine anxiolytic/endovascular association noted above **(Supplementary Table S7)**.

## Discussion

In this multistate claims-based cohort of 54,945 patients with a first observed UIA diagnosis, the prevalence of every assessed mental-health diagnosis was higher in the year after discovery than in the year before, with the largest absolute increases in depression, anxiety, and insomnia or sleep disorder. New psychotropic medication initiation in the year after diagnosis was common, occurring in nearly 1 in 11 eligible patients for benzodiazepines. In adjusted models, surgical clipping was associated with higher odds of initiation across 5 of 6 medication classes compared with untreated observation, and endovascular treatment was associated with higher odds of benzodiazepine, SSRI, and sedative-hypnotic initiation. These findings were consistent across a 30-day diagnostic lag analysis and an alternative SAH/rupture exclusion definition.

These findings are consistent with a growing body of evidence that a UIA diagnosis carries a measurable psychological burden independent of rupture risk. Cross-sectional and case-control studies have linked UIA diagnosis with elevated anxiety and reduced quality of life even without rupture.^1,2,12^ A nationwide South Korean cohort found a higher long-term incidence of mental-illness diagnoses among untreated UIA patients compared with matched non-UIA controls, and a recent multinational registry study linked post-diagnostic anxiety and depression to a reduced likelihood of preventive treatment and worse rupture and mortality outcomes.^5,6^

This study expands on that literature in two ways. First, rather than relying on diagnostic codes alone, we captured a pharmacologic signal, new psychotropic drug initiation and persistence, which reflects treatment-seeking behavior beyond what a recorded diagnosis conveys. Importantly, a comparable signal has been described after aneurysm rupture, where survivors of aneurysmal SAH initiate antidepressants at more than twice the rate of matched controls, suggesting that pharmacologic dispensing is a sensitive marker of psychological burden across the aneurysm-care continuum.^9^ Second, rather than comparing UIA patients with external controls or a single management group, we described diagnosis and medication trajectories across the full spectrum of observation, endovascular treatment, and surgical clipping within one US-insured cohort. This design shows that the increase in mental-health morbidity after UIA discovery reported previously is not confined to observation alone, and that it is measurable in prescribing patterns in addition to diagnostic codes.

The association between surgical clipping and higher psychotropic initiation across nearly every medication class is, to our knowledge, a new observation. This association was robust to adjustment for index year, whereas the corresponding endovascular associations were more sensitive to it. Plausible explanations include perioperative pain, longer hospitalization and recovery associated with craniotomy, and heightened baseline anxiety among patients who elect or are selected for more invasive treatment. Benzodiazepines were both the most commonly initiated medication class and the class with the lowest 1-year persistence, consistent with predominantly short-term or situational use around the time of diagnosis or treatment rather than chronic anxiolytic therapy. Still, roughly 1 in 6 new benzodiazepine initiators met a long-term exposure threshold of at least 120 days within the first year. Given the mean cohort age of 59 years and established concerns about long-term benzodiazepine use in older or medically complex patients, this subgroup may benefit from medication review and counseling regarding non-benzodiazepine or nonpharmacologic approaches to anxiety management at the time of aneurysm diagnosis or treatment.

Taken together, these findings support incorporating brief mental-health screening into UIA care regardless of the chosen management strategy, with particular attention around the time of invasive treatment, when psychotropic initiation was most frequent, and attention to long-term benzodiazepine exposure among new initiators. Multidisciplinary aneurysm clinics that integrate psychological support at the time of diagnosis may be well positioned to address this need.

Several limitations warrant consideration. Diagnosis and medication codes cannot capture symptom severity, confirm medication ingestion, distinguish new psychiatric illness from recognition of preexisting symptoms prompted by increased health care contact after UIA discovery, or establish whether the first observed UIA claim represented the first-ever clinical diagnosis. Observation alone reflected the absence of an observed repair code rather than confirmed clinical surveillance, and aneurysm-specific characteristics associated with rupture risk and treatment selection (eg, size, location, morphology) were not available, so adjusted management-strategy associations may be affected by residual confounding by indication. Finally, MarketScan databases capture a predominantly employer-insured, Medicaid, and Medicare Supplemental population and may not generalize to uninsured or single-payer settings.

## Conclusion

Mental-health diagnoses and psychotropic medication initiation increased after UIA discovery, and medication initiation was most pronounced among patients treated with surgical clipping. These findings support integrating psychological assessment into aneurysm management programs and monitoring for long-term benzodiazepine exposure. They also provide claims-based benchmarks, grounded in a large US population and spanning the full management spectrum of UIA, for future prospective studies of psychiatric outcomes across the UIA care continuum.

## Data Availability

All aggregate data supporting the findings of this study are provided within the article and its Supplementary Material. The patient-level claims data were obtained under license from the Merative MarketScan® Research Databases and cannot be publicly shared or redistributed by the authors because of data-use agreements and commercial licensing restrictions. Qualified researchers may request access directly from Merative (https://www.merative.com/real-world-evidence), subject to its licensing requirements. The authors did not receive special access privileges to these data.

## Acknowledgments

None.

